# Perceptions and Attitudes of Prison Staff towards ADHD – double jeopardy for inmates affected

**DOI:** 10.1101/2020.09.14.20183152

**Authors:** Anna Buadze, Nadine Friedl, Roman Schleifer, Susan Young, Andres Schneeberger, Michael Liebrenz

## Abstract

**Background:** Attention deficit hyperactivity disorder (ADHD) is a neurodevelopmental disorder that is associated with risk-taking behaviors, poor self-control and interpersonal difficulties. Affected individuals have an increased probability of involvement in the criminal justice system, contributing to a higher rate of arrest and imprisonment compared with the general population. Current data on the prevalence of ADHD among prisoners reports rates of 26%, but finds them inadequately treated.

**Aims:** Because prison staff play a central role in the identification of inmates with mental disorders, they could well be key to improving provision of care. There is however little knowledge of the conceptions, perceptions and attitudes of prison staff towards ADHD. Such information could help to identify starting points for awareness training and to further implementation of specific ADHD treatment.

**Methods:** To bridge this gap, we employed a qualitatively driven mixed methods design combining qualitative data collection in the form of narrative interviews with 19 prison staff from a Swiss correctional facility with quantitative data collection in the form of a survey that included the Attitudes Toward Prisoners scale. The interviews were analyzed with QSR NVIVO 11 and a qualitative content analysis approach was used to evaluate findings.

**Results:** Prison staff were generally aware of ADHD and its symptomology, believed it to a be “real”, but “fashionable” disorder and favored hereditary-genetic or biological explanatory models for its development. They viewed inmates with ADHD rather negatively, as complicating correctional efforts, perceived them as sticking out, as tying up more resources and as frequently being involved in confrontations. Our findings suggest that difficulties in pragmatic aspects of communication and language comprehension may be perceived “as not listening or following instructions” creating additional tensions. Consequently, inmates with ADHD are more often exposed to disciplinary sanctions, such as solitary confinement - an intervention deemed “necessary” by staff. Therefore, staff training on ADHD might need to cover evidence on adverse effects. Non-pharmacological interventions for treatment were preferred and considered to be highly efficacious. Skepticism towards pharmacological treatment prevailed, even when benefits from stimulant medication were described. Acceptance of multimodal treatment among prison staff may require customized strategies.

## Introduction

Over the course of the last decades, the established view in the medical literature has regarded attention deficit hyperactivity disorder (ADHD) as a neurodevelopmental disorder that is highly prevalent in both childhood and adolescence and, if left untreated, can be accompanied by severe functional impairments (De Graaf et al., 2008; Faraone et al., 2000; Wilens, Faraone, & Biederman, 2004). Those affected exhibit a triad of inattentiveness, impulsiveness and hyperactivity and show limitations in multiple areas and through all stages of life (Bernardi et al., 2012; Gjervan, Torgersen, Nordahl, & Rasmussen, 2012; Volkow & Swanson, 2013). In comparison to healthy controls, individuals with ADHD not only have a higher rate of conflicts in partnerships, job losses, accidents, traffic offences and substance use disorders, but are also more frequently involved in legal disputes, be they in civil or criminal law (Biederman & Faraone, 2006; Chang, Lichtenstein, D’Onofrio, Sjölander, & Larsson, 2014; Fletcher & Wolfe, 2009; Murphy & Barkley, 1996). The extent of the latter are best exemplified by current studies on the prevalence of ADHD among prisoners, which report rates of 20-40% for incarcerated individuals (Edvinsson, Bingefors, Lindström, & Lewander, 2010; Rasmussen, Almvik, & Levander, 2001; Rösler et al., 2004). A recent meta-analysis pooled 102 original studies from 28 countries published between 1985 and 2017 that included 69,997 individuals living in detention and reported a prevalence rate of ADHD in prison settings of 26.2 % (Baggio et al., 2018). While there has been some debate surrounding the diagnostic accuracy of self-assessment instruments affecting the overall reliability of prevalence rates of ADHD in prisons (Eme & Young, 2017; Murphy & Appelbaum, 2017), the authors furthermore found no significantly different prevalence estimates between studies using screenings for ADHD and those using clinical interviews (Baggio et al., 2018). Compared with the prevalence of ADHD among the general population, the reported rates represent a five-to ten-fold increase for people living in detention (S Young, Moss, Sedgwick, Fridman, & Hodgkins, 2015). On the other hand, there are studies that show that the real possibilities of care for inmates with ADHD are very limited, both in terms of access to diagnostic assessment and initiation of multimodal treatment (Berryessa, 2017; Hall & Myers, 2016; Susan Young, Gudjonsson, et al., 2018). In addition, it has been observed in everyday clinical practice that the continuation of pharmacological therapy for patients treated with stimulants before imprisonment is also extremely complicated and unlikely (Anna Buadze et al., 2020). This lack of care may reflect the challenges faced by those working in the prison system against the background of the comorbidities (conduct disorder, antisocial personality traits or disorder, comorbid substance use disorder) frequently encountered among inmates with ADHD (Hall & Myers, 2016). One point repeatedly mentioned in the literature is the fear of diversion and distribution of prescribed stimulants to other inmates (Appelbaum, 2009). For example, Burns argued for a limit to prescription of stimulants in prisons and named the high prevalence of substance use disorders, the potential of misuse, the possibility of diversion in exchange for money, everyday necessities or other substances, the intimidation of inmates with ADHD to surrender prescribed stimulants to others, security, administrative challenges for prison health staff, direct and indirect costs, the availability of non-stimulant alternatives, the probability of drug-seeking or manipulation to obtain the medication (malingering) and the overwhelming of psychiatric medication management clinics in prison, as the top ten reasons for doing so (Burns, 2009). Indeed, in some jurisdictions, the distinct preference for pharmacologic treatment with nonstimulant medications (e.g., tricyclic antidepressants, bupropion, and venlafaxine) seems to contribute to low treatment numbers and decreased adherence of inmates with ADHD, even when a treatment protocol has been implemented that offers stimulants to those inmates who have failed treatment with one or more nonstimulant agents (Appelbaum, 2009). For example, Appelbaum et al. reported a stimulant treatment prevalence of only 0.7% over a 24month period in a correctional system that had 16,795 potential male candidates for treatment (Appelbaum, 2011). The lack of access to first line pharmacological stimulant treatment for adults with ADHD is all the more surprising as studies on its effects on criminal conviction rates are already very informative. For example a pharmacoepidemiologic study among 25,656 Swedish individuals with a diagnosis of ADHD reported that for those patients receiving stimulant medication, there was a significant reduction of 32% in the criminality rate for males and 41% for females (Lichtenstein et al., 2012). On a different note, the same study did not report the same violence prevention effects for those adults who were prescribed antidepressant medications as an alternative to stimulants (Lichtenstein et al., 2012).

It has long been understood that prison staff plays a central role in the identification and provision of care for inmates with mental disorders in general (Appelbaum, Hickey, & Packer, 2001). In particular, raising awareness of ADHD is considered to be essential, for example, by the Attention Deficit Disorder Association (ADDA) and its ADHD Correctional Health/Justice Work Group or a recently published expert consensus (Susan Young, Gudjonsson, et al., 2018). There is however little knowledge of the conceptions, perceptions and attitudes of prison staff towards ADHD, although this information could help to identify starting points for the development of awareness training and to further the implementation of ADHD specific treatment approaches. For example, it has not been investigated whether prison staff perceive ADHD to be a mental disorder at all, and, if so, endorse biological and medical explanatory models or believe that it can be caused by environmental factors, such as poor parenting, malnutrition or errors during schooling (Matheson et al., 2013; Russell, Moore, & Ford, 2016). Furthermore, prison staff are usually not trained in management strategies of offenders with ADHD, so personal experiences on “what works, and what doesn’t” become highly important when staff interact with inmates affected, for instance when making decisions about disciplinary sanctions. Further, the attitudes and experiences of prison staff towards treatment in general, and pharmacological (stimulant) and non-pharmacological interventions in particular, is unknown, despite these being important pillars in ADHD care.

Here, we propose to explore these gaps by undertaking a qualitative interview study, which aims to:

- investigate whether prison staff perceive ADHD to be a mental disorder
- explore prison staff beliefs regarding the causes of ADHD
- elaborate on past experiences with inmates with a diagnosis of ADHD
- identify prison staff’s view on the role of therapies in ADHD management during times of detention

## Methods

### 2. Methods

#### 2.1 Study design and reporting

This study employed a qualitatively driven mixed methods design combining qualitative data collection in the form of narrative interviews with quantitative data collection in the form of a survey (Hesse-Biber, 2010) to investigate prison staff attitudes towards mental disorders such as schizophrenia, substance use disorders and ADHD. For the purpose of this article we focus on ADHD and report our findings following consolidated criteria for reporting qualitative research (COREQ) guidelines (Tong, Sainsbury, & Craig, 2007).

#### 2.2 Sampling procedure and setting

A modified site-based approach was used to identify and recruit study participants from Realta prison, located in the Canton of Grisons, Switzerland (Lindlof & Taylor, 2017). It is operated as an “open” correctional facility under regional authority with a capacity of 120 for male prisoners who work outside the prison and have up to 36 hours of leave per week. During their stay in prison it is compulsory for detainees to pursue work activity. A number of working areas are available for this purpose, ranging from a market garden to farming on a cultivated area of 136 ha (336 acres), 300 cattle, 60 dairy cows, 100 large livestock, to a carpenter’s shop, a butcher’s shop and a technical workshop for repairing farming equipment.

Because we expected professional background, function and position, years of employment, gender and age to be important correlates of variation in attitudes and perceptions of prison staff towards mental disorders, these were the characteristics of interest when approaching a “gatekeeper”. The importance of gatekeepers in recruitment of participants from hard to reach communities and complex societies has been described elsewhere (Arcury & Quandt, 1999). In the study at hand, an employee of the Department of Justice, Health and Safety of the Grisons served as the initial point of contact, provided information on the site staff members and helped the researchers to identify individuals who would be appropriate for the study (i.e., had the ability to complete an in-depth interview). In order to minimize possible bias on the part of the gatekeeper in selecting participants, considerable effort was made to ensure recruitment of a sample that incorporated diversity, also in respect to interest in the research topic (or lack thereof). For example, an agreement was reached beforehand that study participation could not only be carried out during regular working hours, but was also remunerated as full working time for participants. The only exclusion criterion was unwillingness to give written informed consent. For logistical reasons recruitment continued past the point where saturation was reached.

#### 2.3 Data collection and interview

To investigate whether prison staff perceive ADHD to be a mental disorder, to explore participants’ beliefs regarding the causes of ADHD, to elaborate on past experiences with inmates with a diagnosis of ADHD and to identify prison staff’s views on the role of therapies in ADHD management during times of detention, we conducted single, semi-structured, in-depth interviews lasting between 25 and 66 min, with an average duration of 51 min. We used a self-developed and flexible interview guide which also covered attitudes towards other mental disorders. Two female researchers (NF and AB) conducted the interviews. NF was at the time a Master’s student at the Faculty of Human Sciences, Institute of Psychology, preparing a thesis under the supervision of ML, a forensic psychiatrist and faculty member of the medical school. AB, who was an attending physician at the Psychiatric University Hospital, Zurich, heading the specialized outpatient clinic for ADHD and with experience in qualitative interviews, trained NF and conducted the initial two interviews in the presence of NF. The following interviews were all conducted by NF with regular feedbacks given by ML based on audiotapes and transcripts.

The research team itself had gathered previous experience in employing qualitative research methodology on perceptions towards ADHD, SUD and psychosis among the general population, medical and legal experts and affected individuals. Results have been reported elsewhere (Ana Buadze et al., 2020; Buadze, Stohler, Schulze, Schaub, & Liebrenz, 2010; Canela, Buadze, Dube, Eich, & Liebrenz, 2017a, 2017b; Canela et al., 2019).

Before the interviews, participants had an understanding that NF had a background in psychology and that the research represented a collaboration between the Department for Justice, Health and Safety, Grisons, and several psychiatric institutions and that the research would address prison staff’s experiences with inmates suffering from a wide variety of mental disorders.

All interviews were conducted in Swiss German, an Alemannic dialect spoken in the German-speaking part of Switzerland and in some bordering Alpine communities. Participants were encouraged to speak this dialect in order to make them feel more comfortable. Open-ended questions and non-leading probes were used to encourage participants to speak freely and to elaborate on their statements. Paraphrasing and summarizing main points during the interviews helped minimize misunderstandings and clarify ambiguous statements. Interviews were – with the exception of the initial two interviews – conducted on a one-to-one basis and were digitally recorded. Field notes were taken afterwards.

By grounding the questions in participants’ practice experiences, and by reformulating the questions, we sought to avoid generalized responses. All interviews took place in an office of the correctional facility. The office was reserved for the interviews to ensure that interviewer and participants were undisturbed. Water was available as a refreshment. There were no repeat interviews.

All subjects provided additional biographical data and provided information in regards to education, work experience and experiences with mental illness in the form of a digital survey. The questions formulated for this purpose resembled a query previously used by Callahan et al. (Callahan, 2004). Additionally, a German version of the Attitudes toward Prisoners (ATP) scale was used to evaluate prison staff’s personal attitudes toward inmates without a mental disorder. The ATP scale was translated from English into Standard German by NF and checked for plausibility and comprehensibility by ML. The original English version of the ATP was developed by Melvin et al. and consists of 36 statements about detainees of prisons, for example: “Prisoners are different from most people”, “You should not expect too much of a prisoner”, “Prisoners are just plain immoral” (Melvin, Gramling, & Gardner, 1985). Staff was then asked to rate these statements with respect to inmates on a 5-point scale ranging from strongly disagree to strongly agree. Total score can range between 0 to 144, with a score of 72 indicating a neutral attitude. The original ATP has a moderate to high internal consistency and test-retest reliability (r= .82). To obtain biographical data and to conduct the ATP, a survey was set up employing REDcap software (Harris et al., 2009) enabling participants to fill in the questionnaire onsite using a mobile device.

#### 2.4 Data analysis

Interviews were digitally recorded using an Olympus DS-7000 voice recorder and then transcribed verbatim into Standard German. Whereas Swiss German is commonly only spoken, Standard German is traditionally used in writing and transcription in Switzerland, which is why all interviews were written down in Standard German using a word processor (Microsoft Word). After removing identifying information, each transcript was assigned a code number. The transcripts were not returned to the participants. Subsequently the transcripts were uploaded into QSR NVIVO 11 for Windows (see below).

The procedure regarding the content analysis differed only slightly in comparison to previous studies by our research group and has been described in detail before (Ana Buadze et al., 2020). Qualitative analysis of the interview data was done independently, initially by NF focusing on SUD, and subsequently for the purpose of this publication by AB and ML. AB and ML analyzed the material blinded as to participant identity. A comparison thematic approach, identifying common and new themes related to the research aims was used. For this research, the interviews were analyzed with QSR NVIVO 11 for Windows, a qualitative data analysis software (QDAS) (Kaefer, Roper, & Sinha, 2015). This software was used to organize the semi-structured interviews, to set up case nodes, to code emerging themes and to visualize the data. Coding centered on identifying common and unique themes related to the research aims, as well as omissions within the interview transcripts.

The coding process ensured a systematic, comprehensive and detailed reading of each interview transcript. First, the coders familiarized themselves with the transcripts in order to identify the different subjects of interest. After several interviews had been coded, the categories for the study were redefined, reviewed and revised in a consensual manner at meetings between AB and ML. When there was disagreement regarding the coded material, ML applied the final code. As a result of the coding process and for the purpose of this paper, four main categories were identified and selected: a) personal stance towards ADHD, b) explanatory models, c) experiences with previously diagnosed inmates, and d) attitudes towards necessary interventions. An overview of the categories is shown in Fig 1.

**Figure 1:**
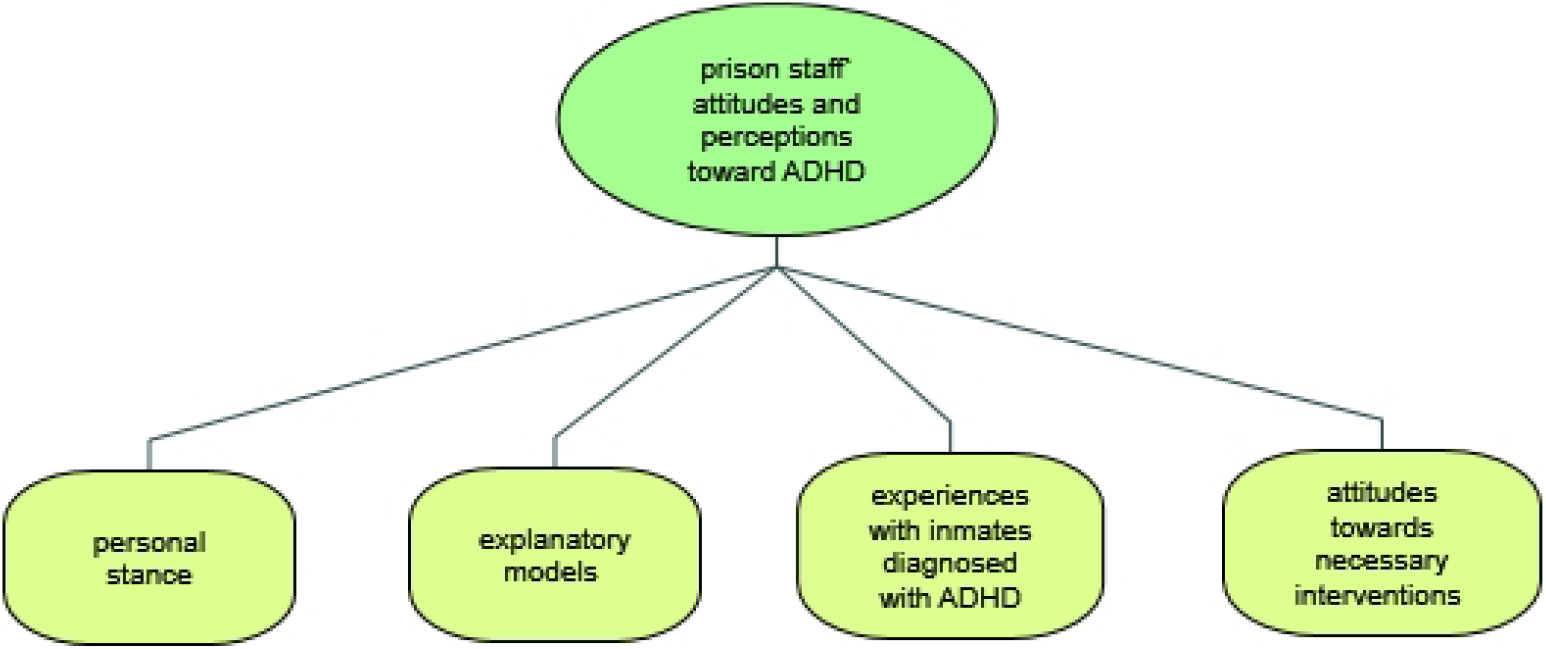
Main categories of prison staff attitudes and perceptions towards ADHD

To illustrate the categories and for reporting purposes, examples of coded quotations were chosen by AB and ML and translated from German into English by ML. Deepl Translator a machine translation service launched in August 2017 was used to support and simplify this translation process. Quotations were then improved by a bilingual German/English speaker (ML) and edited by an English native speaker (Heather Murray) to ensure readability for an international audience.

The quantitative sociodemographic data were evaluated using SPSS version 24. Attitudes towards prisoners with and without a mental disorder were compared using paired sample t-tests and Bonferroni correction (p<.00138).

The research was conducted in accordance with the 1964 Helsinki Declaration and was authorized by the independent ethics committee (IEC) of the faculty of Human Sciences of the University of Bern. All participants were assured confidentiality, and gave their written informed consent for the study and, specifically, for the digital recordings of the interviews.

## Results

### Sample descriptions

During this study the research team established contact with 21 subjects. Of these, one declined to participate. The barrier to participation for this potential participant could not be determined. Another potential subject who had initially agreed to participate was impeded due to an unexpected illness. The scheduling of the interviews proved to be complex, as the employee’s absences and changes in work shifts needed to be considered. Despite careful planning, several appointments had to be rescheduled because of unexpected changes in the shift plan, which however did not lead to reduced participation. On the contrary, subjects showed great interest in the research topics. In total, 19 subjects provided their written, informed consent. All completed the interview. None of the participants withdrew their consent at a later time.

The final sample (n=19) was composed of a higher percentage of male staff (73.3%) than of females (26.3%). The mean age of the participants was 50.4 years (± 10.6 years) with a mean of 15.9 years (± 11.6 years) of working experience in the correctional system. The vast majority of participants (84.2%) had completed vocational training outside the prison system and received a federal diploma before starting to train as correctional officers, as this is a prerequisite for an application to justice department authorities in most cantons (Rohrer & Trampusch, 2011). As intended, the professional backgrounds of the participants reflected great diversity and ranged from training as a farmer, forester, agricultural machinery mechanic, head cabinetmaker, nurse, surveyor to cook, bricklayer and precision mechanic. Regarding their specific correctional training, 94.7% of participants stated that mental disorders were covered in some from, at least theoretically during courses. A total of 72.2% judged their training on this topic to be sufficient for their everyday professional life. On the other hand, only 52.6% reported having been specifically trained to work with people in detention suffering from mental disorders, while a further 30% held the belief that the training they had received was insufficient to prepare them for situations they had encountered while working in the correctional facilities. The majority of participants had had experience with mental disorders in their private environment. Over 70% knew someone who had consulted a psychiatrist or a clinical psychologist or who had been treated as an inpatient in a mental health institution. A further 15.6% believed they suffered or had suffered from a mental disorder themselves. More detailed baseline demographics of participants are illustrated in Table 1.

**Table 1.** Baseline demographics of participants

| Sociodemographic variables | n=19 |
| --- | --- |
| Age (mean±SD) | 50.4 ± 10.6 |
| Gender, male (n, %) | 14 (73.7) |
| Nationality, Swiss (n, %) | 17 (89.5) |
| Years working in the correctional system (mean±SD) | 15.9 ± 11.6 |
| Highest education (n, %) |  |
| Apprenticeship | 7 (36.8) |
| Higher vocational school | 9 (47.4) |
| University degree | 3 (15.8) |
| In possession of a vocational and education training diploma, yes (n, %) | 16 (84.2) |
| Career training as a correctional officer started/graduated, yes (n, %) | 10 (52.6) |
| Some knowledge about mental disorders was imparted during training, yes (n, %) | 18 (94.7) |
| Caring for people in detention with a mental disorder practiced during training, yes (n, %) | 10 (52.6) |
| Knowing someone who consulted with a psychologist/psychiatrist, yes (n, %) | 15 (78.9) |
| Knowing someone who sought treatment in an inpatient psychiatric hospital, yes (n, %) | 14 (73.7) |
| An acquaintance | 5 (26.3) |
| A friend | 2 (10.5) |
| A family member | 5 (26.3) |
| A relative | 2 (10.5) |
| Have/ had the feeling of suffering from a mental illness themselves, yes (n, %) | 3 (15.8) |

### Quantitative Research findings

As mentioned in section 2.3, the ATP questionnaire was used to gather the data related to the participants’ attitudes towards inmates. The possible scores of the ATP questionnaire can range from 0-to-144. An individual with a low score views offenders negatively, as deviant and as incapable of rehabilitation and/or of positive change, whereas a high score is indicative of a more favorable positive attitude towards inmates. All interviewees completed the ATP questionnaire. The mean total ATP score of this sample was 90.58 (SD= 16.804).

### Qualitative Research findings

In qualitative research, saturation is commonly defined as the point when no new themes arise and was achieved in this study after 15 interviews. Because additional interviews were agreed upon prior to reaching saturation, these were followed through with, further broadening the spectrum of individual responses.

#### Participants’ general perception of ADHD

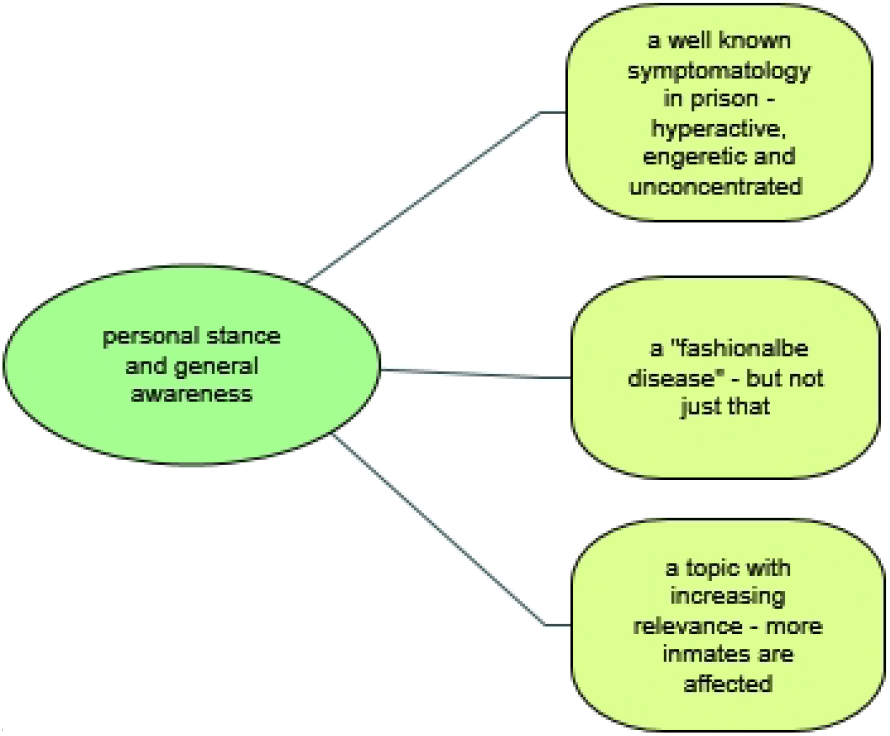

#### A well-known symptomatology in prison: hyperactive, energetic and unconcentrated

Unexpectedly and across all occupational groups, all participants had heard about the disorder and/or had had personal experience with inmates with ADHD, expressing themselves in a complex way on the topic. Furthermore, some of the participants interviewed by us mentioned ADHD or ADHD-like symptomatology in the context of their general and/or opening remarks on how to deal with detainees with mental disorders. It became clear that the disorder and its resulting effects on everyday prison life had reached the general consciousness of the prison staff. This assessment can be exemplified by the statement of one staff member who remarked that he “did not know that ADHD existed in adulthood until he started working in prison.” The description of the symptomatology, although presented by laypersons, deviated only in nuances from the terminology used in medical literature and revolved around terms such as “high-energy”, “explosive”, “hyperactive”, “unfocused” and unconcentrated.”

> *Yes, what strikes me in the prison is that there are always psychologically remarkable inmates who cannot keep to the rhythm of everyday life. For example, I didn’t know there were, uh, adults with ADHD. I didn’t know that. And when you (meet) such prisoners later on, they attract attention when they are so hyperactive. But I did not know until then that this was due to ADHD. I didn’t understand it, until I came to work here*.
>
> *ID 9*

#### A “fashionable” disorder – but not just that

In view of the reservations described in the literature about ADHD, it was important for us to depict in particular the general attitude and any skepticism of prison staff. For this purpose, we used follow-up questions that dealt with the characterization of ADHD as a “fashion diagnosis”, hence a diagnosis that is currently enjoying particular popularity. While participants considered the disorder to be “fashionable”, similar to the diagnosis of “burn-out”, the majority were of the opinion that the “phenomenon” existed, i.e. that there was a “true core” behind it. The interviewees considered the “accuracy” of expert diagnosis as the basis for initiating treatment to be a major problem; they thought that experts were not able to accurately differentiate between those who are impaired but “generally o.k.” and those requiring pharmaceutical treatment. In this context, comparisons to “past-times” were repeatedly made, arguing that, today, all those who could not “function” received a diagnosis, whereas “nervous-fidgety” behavior would just have been accepted by society in the past. Although these statements evoke the idea of “overdiagnosis”, such explicit wording was not used by our participants.

> *Um, I think it’s a really hard disease to diagnose, I strongly believe that. And I actually think that some, um, professionals can’t really diagnose it clearly. But I think this phenomenon exists. It really does exist*.
>
> *ID 8*
>
> *Yeah, it is like “burnout”, it’s fashionable. It’s not easy in the professional world. But when such a hyperventilated (hyperactive, ed.) child tests the limits, I would claim that not everyone has the problem. There are certainly half of those with ADHD who actually have the disease and a dysfunction. But often it is also the case that someone acts out in the school yard. When I act out, I’m accepted, then I’m somebody*.
>
> *ID 10*

#### A topic with increasing relevance – more inmates are affected

Irrespective of reservations about the assessment of ADHD, the disorder was deemed to be a topic of increasing relevance, as more inmates were perceived to be affected and/or diagnosed. Our participants based these perceptions on their many years of experience in the correctional system. In this context, too, it became clear that skepticism about the “existence” of this disorder had generally diminished after joining the prison work force and gaining “hands on” experience.

> *Yes, yes we have ADHD too. Also rising. (…) I think the whole thing will increase in the next few years. This is becoming more and more [common]*.
>
> *ID 13*

#### Participants’ explanatory models

In our sample almost all participants had developed explanatory models about the origin of ADHD. Only two stated that they had no idea about the causes of this disorder and spoke in general terms of “something with childhood”. Although each individual’s etiological concept was somehow unique, it was possible to identify common major themes and shared features. It should be noted, however, that some overlap between themes occurred. The majority of prison staff identified more than one contributing cause and had adopted a multi-factorial explanatory model. We identified three major themes on perceived causation, which we characterize below, starting with the most commonly expressed perception.

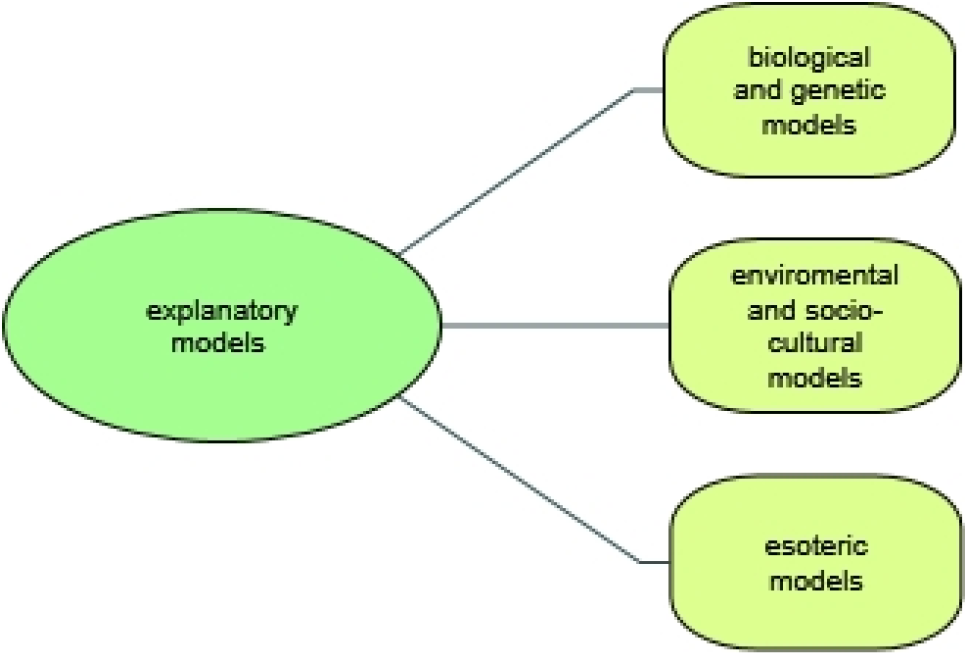

#### Biological and genetic models

Most frequently participants associated the development of ADHD with hereditary-genetic or, more generally speaking, with biological causes, indicating a clear preference for scientific explanations. Occasionally, birth complications (asphyxia) or consequences of alcohol consumption during pregnancy were mentioned.

> *“I think it’s genetic, I think it’s uh biologically phew yes I think genetic and biological above all. So I think the environment has less of an impact. I think the environment has an impact on how someone deals with it. But for the onset of ADHD, that does not arise from bad (…) social influence. Yes genetically, biologically, like that.”*
>
> *ID 15*

#### Environmental and socio-cultural models

Less frequently, prison staff identified environmental, parental or social factors as the main reason for developing ADHD. Factors like increased media consumption, disproportionate use of computer and communication devices, and an overflow in day to day activities were mentioned. The main motive identified was “social pressure” as exemplified below:

> *“I think also familial probably. Their whole youth, their upbringing, school – all around. The whole environment that has developed differently now compared to some years ago, because the pressure from outside is greater and the parents, and many are children of foreigners where this is just one problem and it goes wrong in many other directions. They do not learn German at home and that’s an additional problem. So I think that’s already a big part of the problem.”*
>
> *ID 13*

#### Esoteric models

Mystically explanatory models were adopted by one participant. This explanatory model was linked to a perceived simple solution for symptoms, in this case the consideration of “water veins” and “bed-positioning”.

> *“It may also have been experienced, perhaps even electrosmog in childhood. I could imagine. Or someone always lies over a water pipe as a child, a water vein …, that’s it. I could imagine. The parents are desperate, go to the doctor, get medication everything and, in the end, it would have been only that one had to change the[position of the] bed*”
>
> *ID 11*

#### Participants’ experiences with inmates previously diagnosed with ADHD in a prison setting

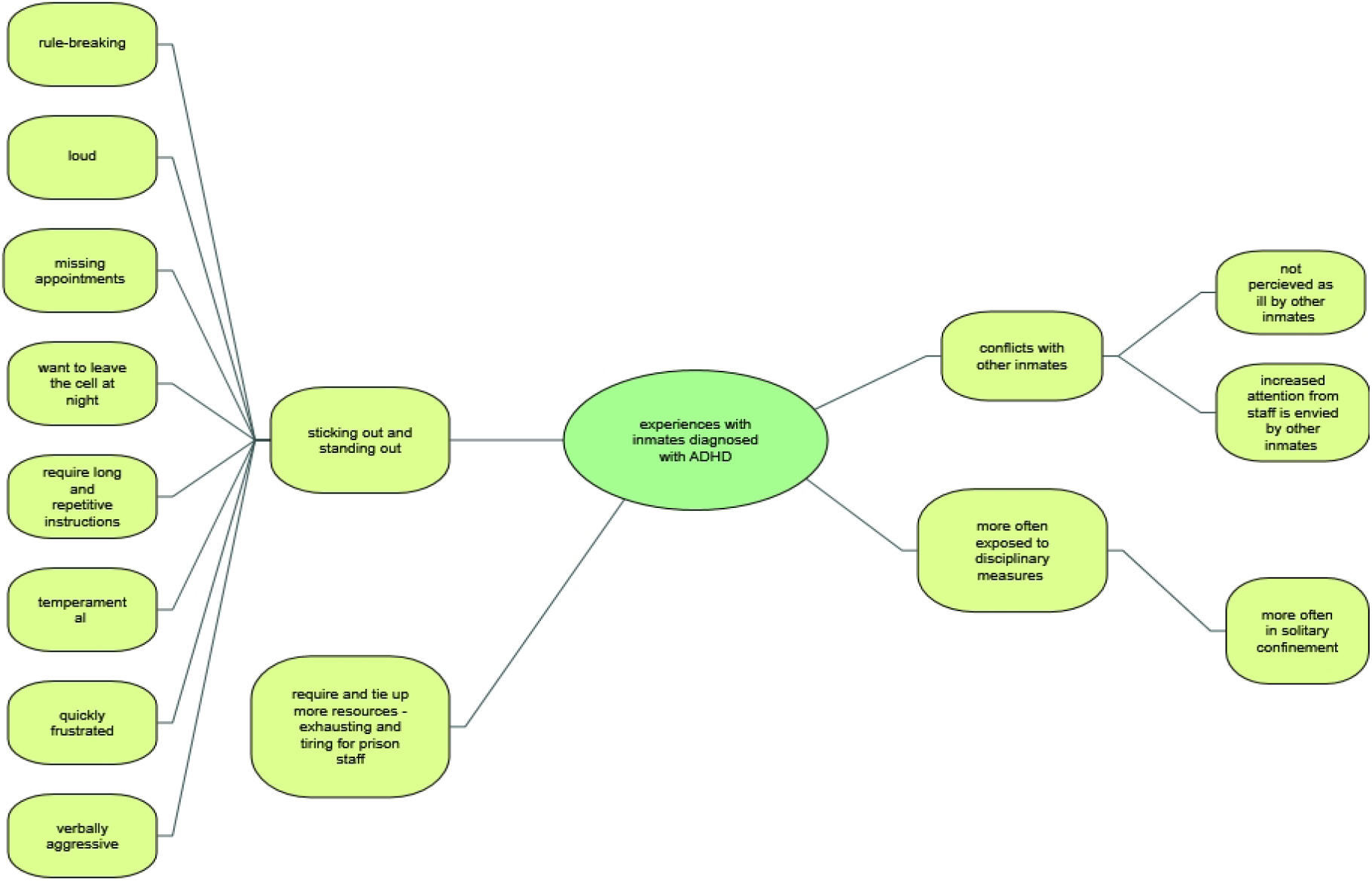

Our participant sample had gathered a great wealth of experience in dealing with detainees diagnosed with ADHD during various stages of incarceration and in a multitude of everyday prison life situations. Although the professional backgrounds of the interviewees differed substantially, and the interactions reflected different areas of responsibility, such as security, vocational rehabilitation or health service provision, views and impressions overlapped considerably and revolved around the same difficulties.

In addition, it must be noted that a majority of prison staff, irrespective of professional background and line of work, seemed to be aware of the mental health status of inmates, indicating a simplified passing on of medical information in this prison. This was justified by the necessities of a strongly labor-oriented correctional facility, in which some jobs assigned to inmates required, for example, the operation of heavy machinery. In this context, it was explained that medical information, such as prescription drug use, must be transparent to most staff.

#### Sticking out and standing out

Analysis identified several important interwoven factors which, in the eyes of prison staff, complicated correctional efforts on an individual level and moreover impacted inmates collectively. Participants had experienced inmates with ADHD as “sticking out and standing out” and perceived them as loud, temperamental, fidgety, quickly frustrated and verbally aggressive. It was commonly reported that this group of detainees were the ones who violated prison rules. In the eyes of the participants, they were the ones who wanted to leave the cell after lock-in, were unorganized and missed appointments, for example, in their work environment or in health services, and did so even when this meant incurring significant disadvantages. Other prison staff indicated that inmates with ADHD required longer and more repetitive instructions, more explanations and generally more attention compared to prisoners who did not have this disorder. In this context, some participants acknowledged different degrees of severity of the disorder, resulting in varying impairments of social functioning.

> *Those who are always tingly, can’t sit quietly, are always on the move – and that’s difficult in the evening when they’re locked up. In the first days these are the ones that constantly ring (the bell) and ask if they can be let out for a moment, because they have to run. But that, uh, yes in the night you have time to explain why it’s not possible and why they have to stay in overnight (…) and that is enough even if you just take the five minutes to quickly talk to them. And then they have the feeling that “I’m not alone”, and at night it’s very quiet in the prison, so it’s possible. Just a few words can help. I have the feeling they need a bit more attention now and then, uh, by the fact that they can get medication from us of course, uh, there is automatically a short conversation and that every now and then I have the feeling they want that, too*.
>
> *ID I*
>
> *The inmate with ADHD just stands out, he simply stands out among all the others. Or if he’s burdened with that. He can’t sit still and things just don’t work out. Always compared to those who do not have the syndrome (…): Yes, and that they have no patience. Or that they overreact at work as well and also their temperament, I think that they are more quickly frustrated and become aggressive*.
>
> *ID 9*

#### Conflicts with other inmates

A majority of prison staff suggested that inmates with ADHD often came into conflict with other inmates. Surprisingly, this was not only perceived to be the result of impulsive-uninhibited behavior targeted directly at other prisoners, e.g. in the sense of verbal or physical assaults, but was seen as a consequence of the increased social attention given by prison staff, which is envied by other inmates. Because inmates with ADHD demanded longer explanations, this time was then not available to other detainees. Unlike other more obvious debilitating mental disorders, such as schizophrenia, prisoners with ADHD are not perceived as mentally ill by fellow inmates, which is why behavioral abnormalities are less accepted and do not evoke sympathy.

> *This is noticeable among prisoners with ADHD. The other prisoners, the collective body, see these prisoners less as ill. If someone has schizophrenia and is clearly behaving strangely, then the illness aspect is not questioned among inmates. But someone who takes up a lot of time, for example from the foreman, in consulting, that is seen critically. We have office hours and people can come up to the floor and, when the caregiver is free, they can come into the office and chat. You just take turns. And sometimes there are remarks to those with ADHD: “Oh you again, you have a torrent of words today”. And in fact, we really need more time with them. And sometimes this really causes friction among the detainees*.
>
> *I would actually say that these are not the ones that are considered sick. They are rather those who are considered to be buzzing and noisy, always scrounging and such*.
>
> *ID 8*
>
> *Yes, they’re very, very, very difficult, how shall I say, to handle. They talk too much, they are, uh, restless and that makes it very difficult for the others to accept them as they are. I think it’s generally difficult for these men in a community (laughs). I think*.
>
> *ID 12*

#### Require and tie up more resources

Almost all respondents experienced the care of detainees with ADHD as labor-and time-intensive, energy-consuming and generally exhausting. While it was perceived as very important to establish structure in day-to-day routines and a schedule that is both organized and predictable, it was exactly those things that were found to be most challenging to implement, when dealing with this group of inmates. In this context it was often mentioned that inmates with ADHD were easily able to upset carefully crafted prior schedules, if these behaviors were not monitored and managed well.

> *Yeah, um, I’d say it’s the hustle and bustle. They are so difficult to handle, um, because you have to structure them well from the outside, because otherwise it will roll over you or make an hour out of 10 minutes*.
>
> *In order to deal with them, as an advisor or also in the security service, it is necessary to give really clear instructions. Make straightforward announcements, try to put things in order. Especially when the inmate talks a lot. Ask what’s important now. They can be very big energy guzzlers. They can turn all your work upside down because everything is so insanely important, right now*.
>
> *ID 8*

#### More often exposed to disciplinary measurements

Security staff in particular described in great detail the inability of inmates with ADHD to regulate themselves and to stop impulsive behavior, which quickly manifested itself in verbal lapses in times of frustration. In the eyes of security staff, this results in more disciplinary sanctions being imposed on inmates with ADHD, such as placement in solitary confinement. A few of the staff had noticed that such a massive restriction of freedom of movement among this group of prisoners only increased undesirable behavior, but deemed this to be a necessity in order to enforce prison rules and to ensure equal treatment for other inmates. In this context, some staff also indicated that, for those placed in solitary confinement, psychiatric support was not always available quickly enough.

> *Yes, very time-consuming. ’Cause this could go well for a moment, and then they’ll be back somewhere up on the roof. You have to get them back down and if it goes well, everything is fine, but then they do something stupid like using the word “asshole” and they are back in solitary confinement. Naturally that is not possible. That’s not possible for others either. Then they are back in confinement and problems are inevitable. They want to get out, they want to smoke, they want “grass”, they want this and they want that, they are constantly on the “bell” yes, yes, it is time-consuming*.
>
> *ID 13*
>
> *Now, while we are talking, one’s in solitary confinement for five or six days and he’s already in trouble. We have to move him to another facility now, because it’s no longer possible for us to keep him here. He also goes from zero to 100 in no time and won’t calm down. He’s a real “Ritalin-boy”, too. He really needs to be seen by a psychiatrist, but he is only here for 20 days and nobody is going to yank out a leg („bust a gut”) over this. He will be in solitary confinement for another 10 days and then he will be released and we will not hear from him again for one or two years. Then he might return. If they are here for such a short term, there is no chance that a psychiatrist will see them. No chance*.
>
> *ID 13*

#### Attitudes towards necessary intervention

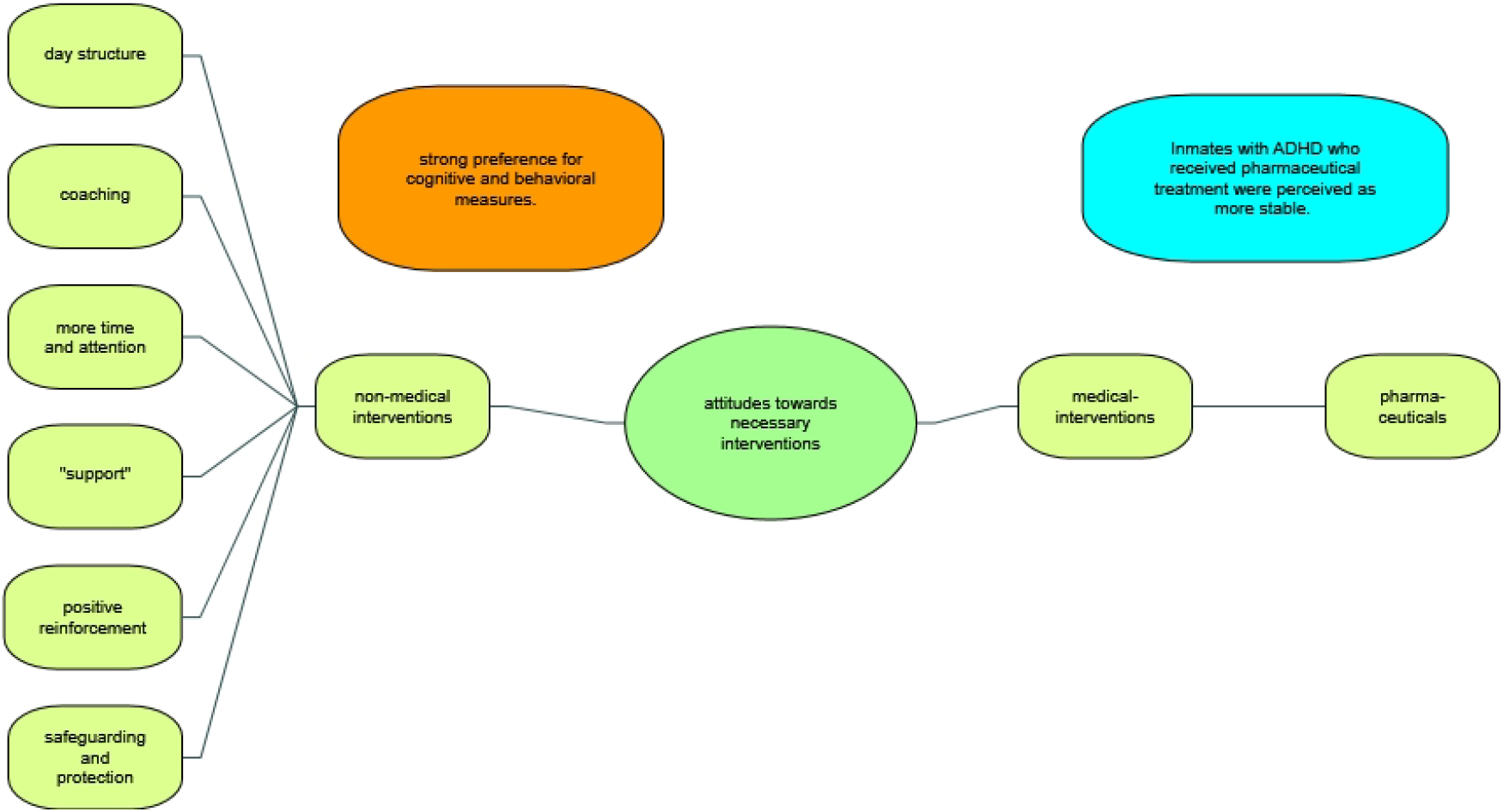

The vast majority of our sample were of the opinion that dealing with inmates with ADHD required a specific approach and had, over the years, themselves developed a variety of interventions that they deemed useful. Regarding treatment for ADHD, almost all interviewees expressed a strong preference for psycho-social interventions, behavioral measures and cognitive training.

#### Non medical interventions

Prison staff specifically mentioned coaching, providing structure, positive reinforcement, time and support when asked about necessary interventions. The meaning of support was interpreted heterogeneously. For example, one staff member reported that he specifically tried to fatigue prisoners with ADHD by pushing them to their physical limits. Interestingly, some staff also found that necessary interventions consisted of safeguarding and protecting affected inmates, and suggested establishing a specific ward for inmates with mental disorders.

> *So I think, uh, interventions gonna have to take place more on the behavioral level. So they need clear structures, clear instructions, no arguing. So I think that is important with ADHD, for people with ADHD. (…) And today I think it’s possible to change something, but it’s, uh, I think a very big effort, in self-control. And although I think that a drug can be helpful, first and foremost it should be about cognitive and behavioral measures. In my eyes*.
>
> *ID 15*
>
> *With me it’s good for them, because when they work with me, they can really give it a go. If I know that one of the detainees has ADHD, then I give him a job that normally two people do. “Put the three stones on the bus.” And if he can do that, that is a form of intervention (medication) without him knowing or realizing it. But that he is exhausted in the evening, that he is needed, that is also healthy*.
>
> *ID 10*

#### Medical interventions

Although inmates with ADHD who were prescribed stimulants such as methylphenidate were perceived as more stable, easier to live with and better manageable for staff, almost all participants showed a certain reluctance towards the necessity of pharmaceutical interventions. None of them was completely against stimulant treatment; however, they clearly did not see it as a viable first-line treatment.

> *Ritalin. If this is well adjusted and they get it, then, uh, they work fine in here. But they’re a little more demanding, I’d say on the average. So they need more attention from us and if they have the medication of course, then it’s easy, then it’s possible*.
>
> *ID 17*
>
> *Some of them are in need of protection. Sometimes you have to, uh, practice with them to get through everyday life here. Help them to acquire knowledge about the structures so that they can learn to move here. Um, they need treatment in the sense that they are medicated, that they are also under psychiatric care and observation, this support has to be guaranteed, so that changes are noticed*.
>
> *ID 8*

## Discussion

In the present qualitatively driven mixed methods study, we investigated the general view and awareness of ADHD held by Swiss prison staff, explored their beliefs regarding causes of this disorder, inquired about experiences with inmates with a diagnosis of ADHD and identified attitudes towards ADHD management during times of detention.

Our qualitative findings on the subject of ADHD should be interpreted in the light of this sample’s fairly negative attitude towards offenders in general, as measured by scores on the ATP. The mean score of prison staff participating in this study was 90.58 (SD= 16.804), which is very similar to scores reported for correctional officers from the USA (90.7), Norway (90) and the United Kingdom, but substantially lower than, for example, scores reported for graduate students in psychology working in the Alabama prison project (113.2) (Hogue, 1993; Kjelsberg, Skoglund, & Rustad, 2007; Melvin et al., 1985). It is therefore not a sample that distinguishes itself by a particularly positive or optimistic attitude towards offenders and their potential for rehabilitation, but it is comparable in attitude to other correctional personnel from jurisdictions with a more punitive approach (Whitman, 2003). This result is surprising for two reasons. Firstly, the sample includes staff from a semi-open prison, which traditionally has a strong rehabilitative character within the Swiss correctional system, and secondly, the majority of the participants were not solely focused on security operations, as employees in this area are known to have more negative perceptions (Kjelsberg et al., 2007; Kury, 2019).

Our results indicate that prison staff working closely with inmates during different stages of incarceration and in a multitude of prison life situations are aware of ADHD and – very specifically -its core symptomatology. In fact, ADHD as such, is an actively discussed topic among prison staff with various professional backgrounds and believed to be of increasing relevance. Interestingly, the skepticism of staff towards the existence of a “Fidgety Phil Syndrome” and the classification of this form of “behavioral abnormalities” as a mental disorder decreased when they started to work in a prison environment, a finding that even held true for individuals who had previously gained experience in general psychiatry. Several explanations for this finding seem plausible: Firstly, it may be simply more likely for staff to encounter individuals with ADHD in a correctional facility, considering that the prevalence of individuals with ADHD symptoms is higher by a factor of 5-10 compared to the general population (S Young et al., 2015). Secondly, individuals with ADHD who are detained in a prison are likely to be part of a subgroup of affected individuals whose functional capacity (as conceptualized by the WHO) is particularly impaired, through a combination of biological, social, personal, and environmental factors (Bölte et al., 2018; Mahdi et al., 2018; Susan Young et al., 2009). It is thereby well understood that people with ADHD who are detained suffer, for example, to a high degree from comorbidities (Rösler et al., 2004; Rösler, Retz, Yaqoobi, Burg, & Retz-Junginger, 2009). Existing symptoms and impairments may thus initially appear to be more pronounced, especially against the backdrop of the highly structured and demanding prison environment with strict schedules, little flexibility and few possibilities to apply self-developed skills and compensation strategies, such as motoric (increased physical activity) or organizational strategies (delegation of tasks, use of electronic devices) (Canela et al., 2017b). Executive function deficits may thus be easier to recognize even for laypersons with limited training (Brown, 2009). Further, it is conceivable that other neurodevelopmental disorders (NDD), such as Fetal Alcohol Syndrome (FAS) and Autism spectrum disorder (ASD), which still receive little attention in the Swiss correctional system and have a substantial rate of overlap, are currently being (mis-) labeled as “ADHD” by staff (Billstedt, Anckarsäter, Wallinius, & Hofvander, 2017; Popova, Lange, Bekmuradov, Mihic, & Rehm, 2011; Susan Young, González, et al., 2018).

With regard to a previous study among the general population from neighboring Germany, it can be said that our qualitative findings complement quantitative results. Speerforck et al. reported recently that more than 90% of the German general population had heard of the terms “attention deficit hyperactivity disorder” or “ADHD,” and 77% of those believed adult ADHD to be a real disease (Speerforck et al., 2019). In addition, a study from the USA concluded that 78% of those who had heard of ADHD believed it to be a real disease, with women and individuals with a higher socioeconomic background most often endorsing this belief (McLeod, Fettes, Jensen, Pescosolido, & Martin, 2007).

Apart from the general awareness and a shared belief that ADHD is a “real disease”, prison staff were under the impression that there was a lack of reliability in assessment of ADHD by experts, especially in respect to the threshold for initiation of pharmaceutical treatment. Furthermore, participants labeled ADHD as a particular “fashionable” diagnosis, i.e., as a disorder that is “popular” for mental health experts to diagnose or for patients to receive. Repeated statements that “behavioral abnormalities were socially more acceptable in the past” and that today “it is simply a matter of making a diagnosis” can also be regarded as indicative for the belief that ADHD is inappropriately mis- and overdiagnosed. Although, according to the knowledge of the authors, there are no studies that have investigated this phenomenon quantitatively or qualitatively among prison staff, quantitative studies among other populations show that this belief is widespread. For example, a survey of Australian general public attitudes towards the acceptability of pharmaceutical treatment for ADHD found that 78% of participants held the belief that individuals are diagnosed with ADHD when they do not actually have the disorder, a stark contrast to current evidence indicating that the majority of patients are underdiagnosed and undertreated (Ginsberg, Quintero, Anand, Casillas, & Upadhyaya, 2014; Hamed, Kauer, & Stevens, 2015; Partridge, Lucke, & Hall, 2014).

Given the perception that ADHD is a “fashionable diagnosis” and a convenient social label, it was surprising that prison staff favored hereditary-genetic or biological explanatory models for the development of ADHD. Environmental and social-cultural models revolving around external factors such as parental and familial stressors, social pressure and critical life events or disproportionate use of computer and communication devices played a lesser role in this sample. Mystical explanatory models were a rarity. The strong emphasis on the biological component found in this sample is also surprising in comparison to other studies investigating explanatory models for ADHD. For example, a study among general practitioners, i.e. a far more specialized clientele, identified factors which were mainly social, parental, environmental but much less hereditary-genetic or biological in character (Shaw, Wagner, Eastwood, & Mitchell, 2003). A representative population survey in Germany (N = 1,008) reported stressful life events, a pressure to perform and problems with parents or family as the top ranked causal beliefs among the German general population for adult ADHD (Speerforck et al., 2019). Furthermore, the authors of this study reported that those individuals who favored a “biogenetic” explanatory model were more open towards treatment by a psychiatrist, psychotherapist and the use of medication (Speerforck et al., 2019). There were some indications that these assertions held true in our sample of prison staff as well. However even those who had adopted hereditary-genetic or biological explanatory models and were in favor of some form of treatment intervention (see below), viewed psychotherapy as the first and pharmaceutical interventions only as a second line or last resort treatment.

Apart from the variance in explanatory models, it became apparent that prison staff, irrespective of professional background, generally viewed inmates with ADHD rather negatively, as complicating correctional efforts on an individual level and for inmates collectively. A recurring motive in this context was that this group of inmates is always “standing out and sticking out”, is quickly frustrated, verbally aggressive, disorganized and violating prison rules. More positive descriptions and attributions that can be found especially in lay literature, such as ADHD as a “gift” and as a disorder related to an increase in “creativity, ingenuity, spontaneity and brightness” were almost absent and largely uncommon (Armstrong, 2010; Coghill, 2005). This may be due to the fact that such attributes are generally not in demand and perceived as counterproductive in a correctional environment (Duguid, 2000).

In the literature there is some anecdotal evidence that inmates with ADHD may become model prisoners during times of incarceration, as it is believed that they benefit from highly structured surroundings (Berryessa, 2017; Hurley & Eme, 2004). Such assertations were not made by the staff we interviewed for this study, although some of the affected offenders had achieved a comparatively high level of functioning. Rather, a majority of prison staff were under the impression that inmates with ADHD more often came into conflict and confrontation with other prisoners. While this is in line with earlier reports from Young et al., who found that inmates with ADHD are six times more likely to engage in physical confrontations with other detainees, for example, as a result of impulsivity deficits leading to verbal- and physical aggression, the statements of some interviewees in the present study show a more detailed understanding of which impairments associated with ADHD increase the likelihood of involvement in critical incidents. For example, it was repeatedly stated by staff that excessive talking and the need for lengthy explanations tied up time that was not available to other detainees and thus gave rise to frustrations among them. Indeed, there is an abundance of literature linking ADHD and difficulties in pragmatic aspects of communication, such as speaking without thinking, interrupting others’ speech or conversations and talking excessively (Bellani, Moretti, Perlini, & Brambilla, 2011; Kim & Kaiser, 2000). Additionally inattentive symptoms have been linked with language comprehension difficulties, a factor that often is perceived “as not listening or following instructions”(Baker & Cantwell, 1992). It seems plausible that these impairments and the resulting difficulties create additional tension between inmates, especially when one considers that prison staff voiced the opinion that inmates with ADHD are not perceived as mentally ill by fellow inmates and behavioral abnormalities are less accepted than, e.g., in those suffering from more obviously debilitating mental disorders such as psychosis. This may be an advantage and a disadvantage at the same time. Research on stigma in prison suggests that people in detention labeled as “mentally ill” are at an increased risk of being victimized by offenders who have not been labeled that way and therefore appear to be of lower hierarchic rank (Edwards, 2000). Inmates with ADHD might therefore not be affected by such direct forms of stigmatization and victimization. This should be a subject of further research. However, disorder-related difficulties are not perceived as such, which is why other inmates show no understanding when breaking prison rules due to symptoms of hyperactivity, impulsivity or inattention is not sanctioned by staff, even if certain members of prison staff are aware of “cause and effect” in relation to ADHD. A recurring motive was that inmates with ADHD are more often exposed to disciplinary sanctions, especially in one of its most controversial forms, i.e., placement in solitary confinement (Andersen et al., 2000; Smith, 2006). From their statements in the present study it could be inferred that prison staff had experienced negative consequences of this form of punishment first hand and believed it to be “counterproductive” especially in offenders with ADHD. At the same time correctional officers were generally of the opinion that this form of disciplinary sanction was “necessary to ensure equal treatment”, contrasting starkly with current evidence and available alternatives (Glowa-Kollisch et al., 2016; Haney, 2018). Even though the specific details of solitary confinement differ significantly between jurisdictions and the duration as a disciplinary sanction is generally limited on a cantonal level in Switzerland to days, future research should review this aspect not only from a medical-ethical, but also from a legal perspective, because some statements in the current study suggest that solitary confinement is imposed on offenders with ADHD even for smaller infractions such as verbal insults and may last up to weeks (Jositsch, Ege, & Schwarzenegger, 2018; Künzli, Frei, & Spring, 2014). Furthermore it could be argued that confining, restricting and limiting movement and exercise for 23 hours a day for individuals with ADHD can be considered inhuman- and ill-treatment or even a kind of “double-jeopardy” because it has repeatedly been shown that physical activity mitigates ADHD symptoms, increases cognitive performance, improves executive function and helps those affected to manage behavioral symptoms (Archer & Kostrzewa, 2012; Fritz & O’Connor, 2016; Gapin, Labban, & Etnier, 2011; Suarez-Manzano, Ruiz-Ariza, De La Torre-Cruz, & Martinez-Lopez, 2018). Thus physical activity can be considered a self-prescribed compensation mechanism, that is often recognized as a “skill” by those affected and is one of the few currently accessible treatment options for adults with ADHD in a correctional setting (Canela et al., 2017b). Other authors have repeatedly called for the implementation of ADHD awareness training and workshops in the correctional system (Susan Young, Gudjonsson, et al., 2018). In light of our findings, it may be useful during the development of such courses to consider including material and evidence on the adverse effects of solitary confinement on offenders with ADHD, to which prison staff may be able to relate, based on their own experiences.

It should not go unmentioned that, in the opinion of prison staff, detained individuals with ADHD required a specific form of “management” or “treatment” in everyday prison life and had identified a variety of interventions that they deemed useful, such as coaching, providing structure, positive reinforcement, time and support. This observation underscores that prison staff might be valuable for providing one-on-one skill-building sessions for inmates with ADHD, once adequately trained (Susan Young, Gudjonsson, et al., 2018). However, in line with existing literature on general public attitudes towards the acceptability of behavioral and pharmacological treatments for ADHD, prison staff perceived psycho-social interventions, behavioral measures and cognitive training as more acceptable than medication (McLeod et al., 2007; Partridge, Lucke, & Hall, 2012; Singh, 2003). Non-pharmacological interventions were preferred by staff because they were believed to be efficacious, resulting in sustainable improvements and were considered to be free of adverse effects – underlining the gap between evidence on efficacy of nonpharmacological treatment and the impact of adverse events associated with nonpharmacological treatments of ADHD and public opinion (NICE, 2018). The skepticism towards pharmacological treatment remained, even when interviewees’ described benefits from stimulant medication and viewed inmates in treatment as more stable, easier to live with and better manageable. Previously it has been reported that these reservations may reflect diffuse fears of possible side effects or stigma that are not easily modified or corrected (Krain, Kendall, & Power, 2005). Interestingly, this skepticism was not the result of negative experiences with the misuse and diversion of stimulants as outlined by Burns et al. (Burns, 2009), but reflected perceptions more in line with that of the general population (Speerforck et al., 2019). In fact, none of the prison staff interviewed in this study spontaneously expressed any concern related to the top ten reasons described in detail in the introduction section. On the one hand, this could be due to positive experiences with the prescription of other highly regulated psychotropic substances such as methadone, buprenorphine but also diacetylmorphine (Heroin) to inmates suffering from opioid dependence in this correctional facility (Uchtenhagen, 2010) (Liebrenz et al, under review). On the other hand, the risk of misuse and deviation of stimulants by inmates in treatment during times of incarceration might be overrated. In fact, recommendations for the restrictive use of stimulants in prison are rated at the lowest level of evidence quality, being based on clinical experience, descriptive studies, or reports of expert committees (Appelbaum, 2009; Murad, Asi, Alsawas, & Alahdab, 2016). What little data exists on the deviation of stimulants by inmates with ADHD who are being treated for the disorder stems, to the authors’ knowledge, primarily from the US, where standards, principles and conditions of detention differ not insignificantly from, for example, those jurisdictions that adhere to the European Prison Rules as drawn up by the Council of Europe (Ministers, 2006). However, even these figures seem encouraging from our perspective: for example, Appelbaum reported about 116 male inmates with ADHD from the Massachusetts state prison system treated with stimulants between 2005 through 2007 and found that 105 (90.5%) adhered to protocol and showed no misuse of stimulants or other medications (Appelbaum, 2011). Since people living in detention are in many jurisdictions entitled to a standard of care equivalent to that accessible for those in the community, this data, in the authors’ view, does not support the notion of generally prescribing stimulants “only after a failure of a complete trial of one or more nonstimulant agents”, as suggested earlier (Appelbaum, 2009).

Finally, the current standard of care in the form of a multimodal treatment for ADHD was, not surprisingly, unknown to the correctional staff interviewed in this study and remained unmentioned. This observation underlines the importance of disseminating information and knowledge on this disorder to employees of correctional facilities (Susan Young, Gudjonsson, et al., 2018). Since almost all respondents experienced the care of detainees with ADHD as labor- and time-intensive, energy-consuming and generally exhausting, and had adopted the view that inmates suffering from this disorder were in need of specific interventions, support and treatment, we are cautiously optimistic that at least in comparable penal institutions, staff may be open towards the implementation of non-pharmacological treatments, offender psychoeducation and psychological treatment programs such as Reasoning and Rehabilitation 2 ADHD (SJ Young & Ross, 2007). In order to develop the understanding and acceptance of stimulant treatment for inmates with ADHD, it might be helpful to point towards the successes of other pharmaceutical treatments for mental disorders, that were once considered highly controversial, but are generally better accepted today by prison staff (Ana Buadze et al., 2020). In addition, public opinion also seems to be changing regarding the use of psychotropic medications in cases of mental illness, as recently pointed out by Angermeyer et al., who reported that attitudes towards pharmacological treatment have become noticeably more favorable over the last two decades (Angermeyer et al., 2016).

### Limitations

These results need to be considered within the limitations of the investigation. First, because this is a qualitatively driven, mixed methods design study based on a modified site-based approach with a gatekeeper to recruit participants, the findings on the staff’s personal stance towards ADHD cannot be generalized beyond this study sample. However, with regard to the explanatory models identified, experiences with inmates previously diagnosed with ADHD and attitudes towards necessary intervention, the sample represents a group of prison staff with a diverse professional background, with multiple years of experience of working within a correctional system and at different stages of training as correctional officers. With such a composition, it is very likely that this sample is similar to those of other correctional facilities in Switzerland, especially since a completed vocational training is a prerequisite for an application to work with justice authorities in most cantons, and specific correctional training courses are organized on a super-institutional level, for example, by the Swiss Center for Expertise in Prison and Probation (SCEPP). In addition, scores on the ATP scale suggest that this sample’s general attitude towards offenders is comparable to those reported for correctional officers from other European jurisdictions and even some common law countries. Second, there are limitations associated with volunteer bias, to which most studies are also susceptible. In order to minimize possible bias of the gatekeeper in selecting participants, much effort was made to ensure recruitment of a sample that incorporated diversity, also in respect to interest in the research topic. As described in more detail above, study participation was carried out during regular working hours and was also remunerated as full working time for participants. Only one of the potential participants contacted declined to participate. The barrier to participation for this potential participant, however, could not be determined. As a qualitatively driven mixed methods design study, this study was not driven by a theoretical framework. Future studies on this subject could, however, use the insights gained here to pursue more focused research. We also recognize that the results may in part be specific to the Swiss legal and penal system. Nevertheless, the literature indicates that some perceptions and views identified in this study, such as a lack of knowledge on multimodal treatment options for ADHD and the skepticism towards the use of stimulant medication, have also been reported from other countries. Our findings provide several relevant insights into views held by prison staff on ADHD as a mental disorder and on individuals living with this disorder while being detained. Most importantly, our findings are based on prison staff’s own reports identifying a range of experiences. These findings were not limited to predefined experiences, as might occur in a survey-based research. Furthermore, a written survey might have increased the likelihood of socially desirable responses.

## Conclusions

This research extends our understanding of members of prison staff’s perceptions and explanatory models of ADHD, their experiences with inmates diagnosed with this highly prevalent neurodevelopmental disorder and their attitudes towards ADHD management during times of detention. Our results indicate that prison staff working with inmates during different stages of incarceration are aware of ADHD and its core symptomatology and believe it to a be “real disorder.” Simultaneously, participants labeled ADHD as a particularly “fashionable” diagnosis and thought that ADHD was inappropriately mis- and overdiagnosed. Unlike other populations, this sample favored hereditary-genetic or biological explanatory models for the development of ADHD over environmental and social-cultural explanations. Irrespective of professional background, staff generally viewed inmates with ADHD rather negatively, as complicating correctional efforts both on an individual level and for inmates collectively, and had had the experience that they more often came into confrontation with other prisoners. While this is in line with earlier reports, our findings suggest that this not only due to physical aggression, but also to difficulties in pragmatic aspects of communication. It seems plausible that these impairments create additional tension between inmates, especially when one considers that prison staff voiced the opinion that inmates with ADHD are not perceived as mentally ill by fellow inmates. A recurring motive in the context was that inmates with ADHD are more often exposed to disciplinary sanctions, especially in one of their most controversial forms, i.e. placement in solitary confinement. While prison staff had experienced negative consequences of this form of punishment and believed it to be “counterproductive”, they deemed it to be “necessary to ensure equal treatment”, contrasting starkly with current evidence. In light of our findings, it may be useful during development of ADHD awareness courses to consider including material on the adverse effects of solitary confinement. It should not go unmentioned that, in the opinion of prison staff, detained individuals with ADHD required a specific form of “management” or “treatment” in everyday prison life. Non-pharmacological interventions were preferred because they were believed to be efficacious and were considered to be free of adverse effects - underlining the gap between evidence and public opinion. The skepticism towards pharmacological treatment remained, even when interviewees described benefits from stimulant medication and viewed inmates in treatment as more easily manageable. Interestingly, this was not the result of negative experiences with the misuse and diversion of stimulants, which was not reported. We are carefully optimistic that at least in comparable penal institutions, staff may be open to the implementation of non-pharmacological treatments. In order to develop the acceptance of stimulant treatment for inmates with ADHD, it might be helpful to point towards the successes of other pharmaceutical treatments for mental disorders, that were once considered highly controversial, but are generally better accepted today by prison staff.

## Data Availability

Excerpts of the transcripts relevant to the study are available on substantiated request from the corresponding author.

## Ethics statement

The research was conducted in accordance with the 1964 Helsinki Declaration and was authorized by the independent ethics committee (IEC) of the faculty of Human Sciences of the University of Bern. All participants were assured confidentiality, and gave their written informed consent to participate in the study and, specifically, to digital recording of the interviews.

## Author contributions

AB/NF: Data gathering, evaluation of data. NF: development of topic guide, obtaining ethical approval. RS/SY/AS: Writing, revisions. AB/ML: Conception, development of topic guide, evaluation of data, writing manuscript, revisions.

